## Supplementary figures and images for "Tumor Purity-Related Genes for Predicting the Prognosis and Drug Sensitivity of DLBCL Patients"

### supplementary figure 1

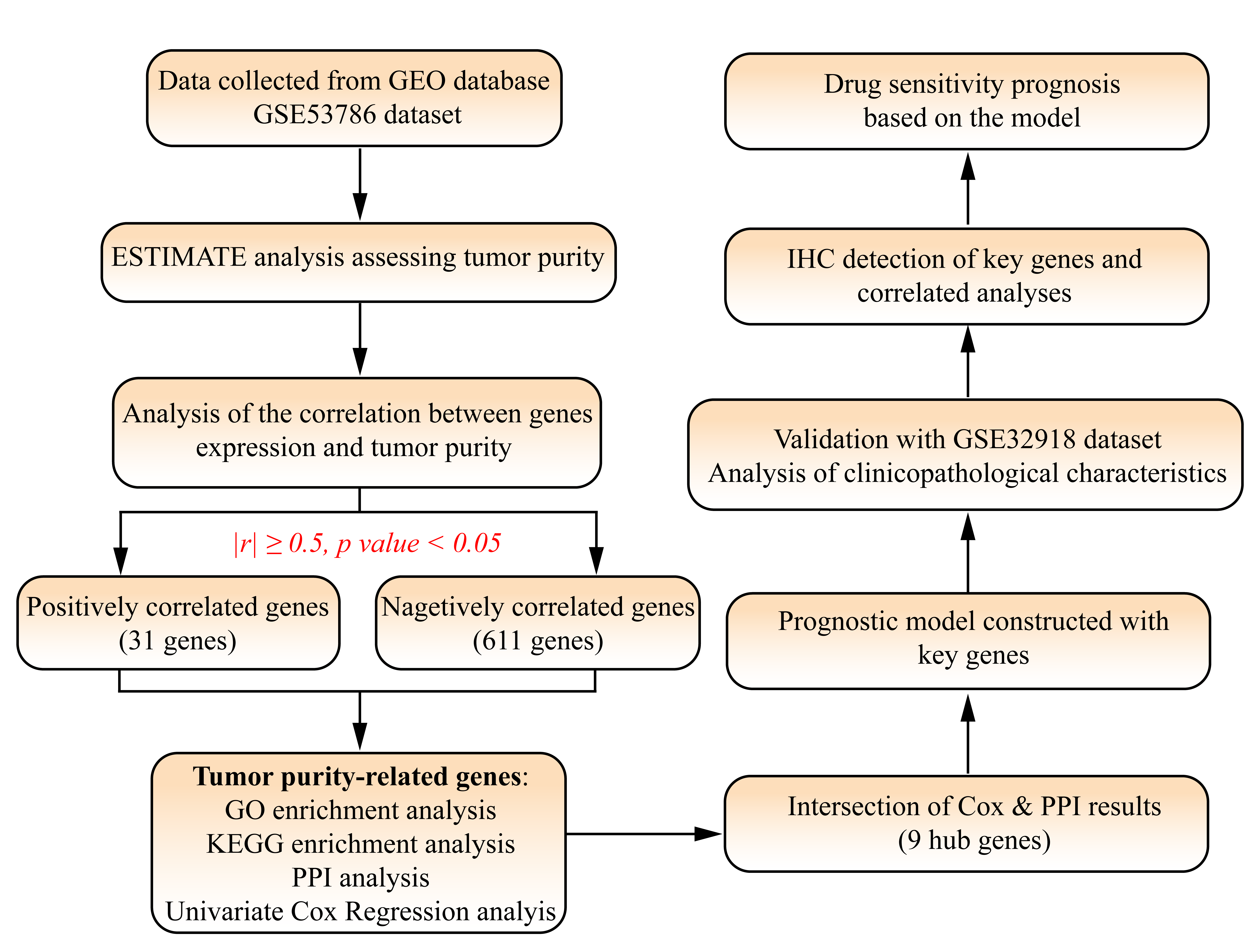

### supplementary figure 2

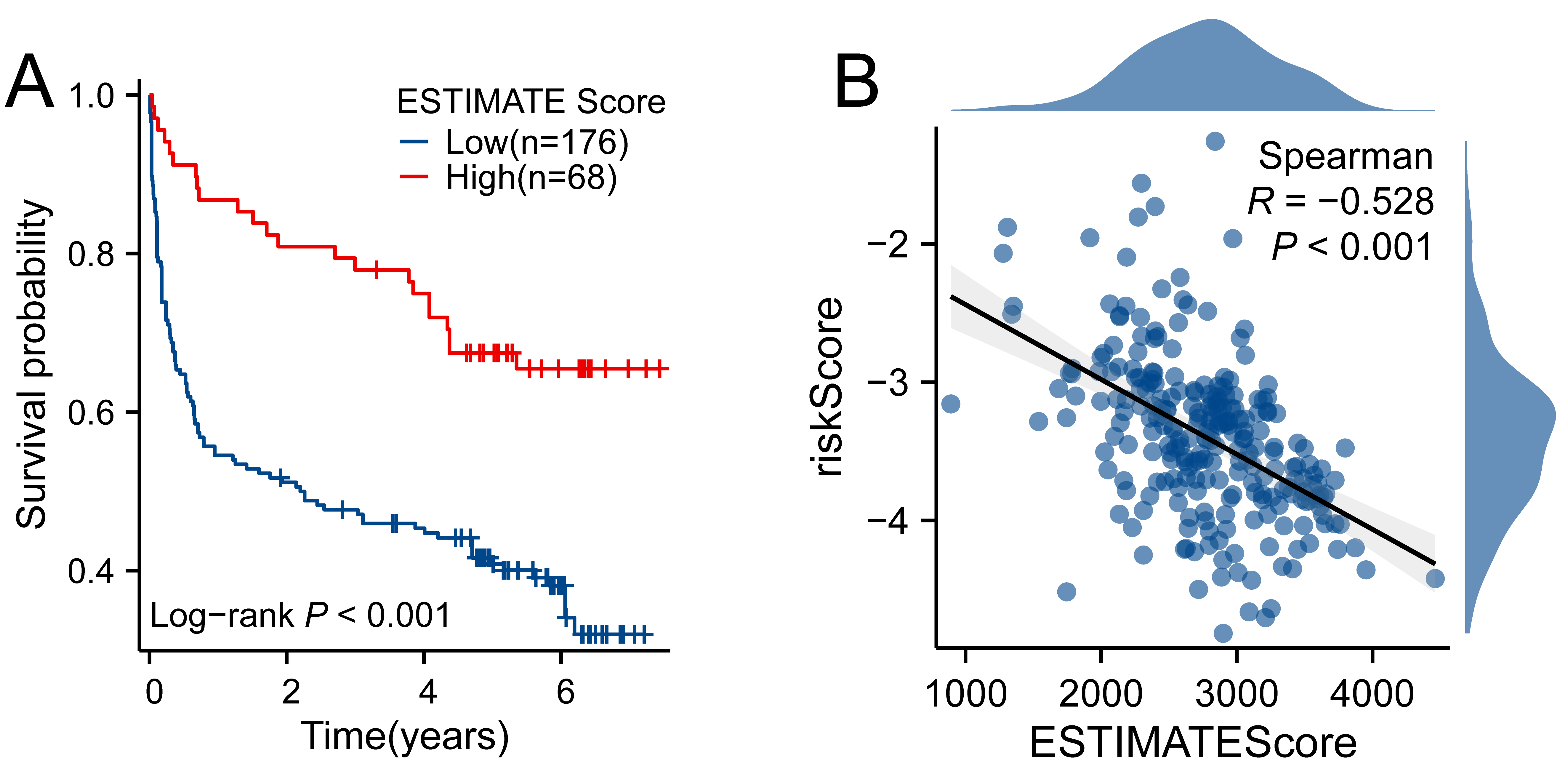

### supplementary figure 3

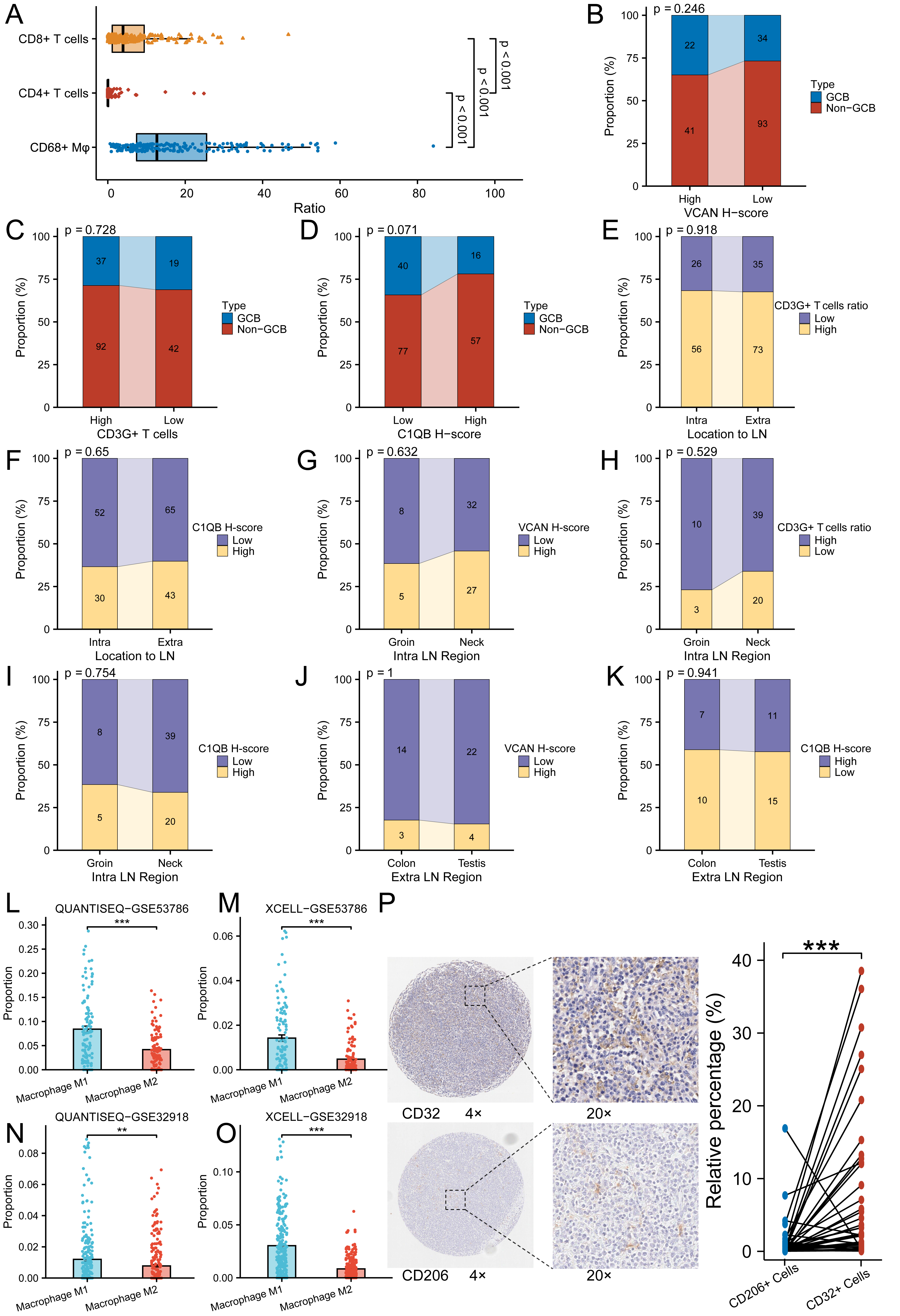
