## supplementary tables for "Tumor Purity-Related Genes for Predicting the Prognosis and Drug Sensitivity of DLBCL Patients"

**Supplementary Table 1 The Clinicopathological Characteristics of CHCAMS Cohort**

| Characteristics | n(%) |
| --- | --- |
| Sex |  |
| male | 106(55.8%) |
| female | 84(44.2%) |
| Age (53.85±1.14^a^) |  |
| ≤60 | 115(60.5%) |
| ＞60 | 75(39.5%) |
| Intra or Extra LN^b^ |  |
| intra | 82(43.2%) |
| extra | 108(56.8%) |
| Position |  |
| neck LN^b^ | 59(31.1%) |
| testis | 26(13.7%) |
| colon | 25(13.2%) |
| groin LN^b^ | 14(7.3%) |
| others | 66(34.7%) |
| VCAN-H Score (cutoff =275.42, 233.37±4.60^a^) |  |
| Low | 127(66.8%) |
| High | 63(33.2%) |
| CD3G+ T cells-ratio (%, cutoff = 2.5%, 14.25±1.32^a^) |  |
| Low | 61(32.1%) |
| High | 129(67.9%) |
| C1QB-H Score (cutoff = 82.41, 65.98±3.01^a^) |  |
| Low | 117(61.6%) |
| High | 73(38.4%) |
| CD68+ Mφ -ratio (%, cutoff = 18.6%, 17.75±1.05^a^) |  |
| Low | 125(65.8%) |
| High | 65(34.2%) |
| CD4+ T cells-ratio (%, cutoff = 0.13%, 0.68±0.20^a^) |  |
| Low | 132(69.5%) |
| High | 58(30.5%) |
| CD8+ T cells-ratio (%, cutoff = 0.375%, 6.69±0.56^a^) |  |
| Low | 24(12.6%) |
| High | 166(87.4%) |

a. mean±SEM, b. Lymph node

**Supplementary Table 2 The prediction of drug sensitivity**

| Drugs | GSE53786 Sensitivity Score (Mean ± SEM) | | | GSE32918 Sensitivity Score (Mean ± SEM) | | |
| --- | --- | --- | --- | --- | --- | --- |
|  | Low | High | P value^1^ | Low | High | P value^1^ |
| Carmustine | 486.997±11.783 | 421.590±10.472 | 0.0000385 | 474.022±8.196 | 432.643±6.370 | 0.000086 |
| Cytarabine | 6.668±0.435 | 5.361±0.307 | 0.028 | 6.894±0.361 | 5.654±0.352 | 0.0000353 |
| Oxaliplatin | 52.990±3.395 | 37.582±1.864 | 9.19×10^-6^ | 48.933±1.720 | 40.808±1.666 | 4.35×10^-6^ |
| Vincristine | 0.203±0.014 | 0.146±0.008 | 0.0005 | 0.219±0.026 | 0.223±0.031 | 0.0047 |
| Vorinostat | 4.509±0.131 | 4.066±0.120 | 0.0138 | 4.618±0.091 | 3.924±0.056 | 7.09×10^-9^ |
| Afuresertib | 13.022±0.581 | 13.420±0.603 | 0.5932 | 13.847±0.362 | 12.380±0.338 | 0.0037 |
| Bortezomib | 0.008±0.0002 | 0.007±0.0002 | 0.0355 | 0.0081±0.0002 | 0.0076±0.0002 | 0.0035 |
| Ibrutinib | 97.082±3.271 | 91.220±3.254 | 0.2068 | 98.641±2.116 | 87.625±1.638 | 0.0000836 |
| Tamoxifen | 37.392±1.129 | 34.720±0.904 | 0.1536 | 37.358±0.627 | 34.545±0.669 | 0.0002 |
| OTX015 | 13.468±0.672 | 11.710±0.485 | 0.0905 | 12.439±0.351 | 12.818±0.590 | 0.1642 |
| Cyclophosphamide | 177.152±3.594 | 170.524±3.350 | 0.203 | 176.726±2.868 | 173.443±3.273 | 0.1019 |
| Dinaciclib | 0.0607±0.0014 | 0.0606±0.0014 | 0.8712 | 0.061±0.001 | 0.065±0.003 | 0.5872 |
| Buparlisib | 2.590±0.049 | 2.617±0.057 | 0.7721 | 2.615±0.039 | 2.643±0.057 | 0.2177 |
| Alisertib | 7.352±0.332 | 6.855±0.354 | 0.1566 | 7.284±0.222 | 6.947±0.247 | 0.1359 |
| Gemcitabine | 0.696±0.111 | 0.595±0.062 | 0.2391 | 0.732±0.104 | 0.818±0.153 | 0.1453 |

1. Wilcoxon rank sum test, P value < 0.05 is regarded as statistically significant.
